# Effects of four-month-long foreign language learning on executive functions and white matter integrity in older adults

**DOI:** 10.1101/2023.09.25.23296063

**Authors:** Giovanna Bubbico, John G. Grundy, Riccardo Navarra, Alessandra Stella Caporale, Chiara Candita, Michela Bouraimis, Miriam Felice, Piero Chiacchiaretta, Alberto Granzotto, Armando Tartaro, Antonio Ferretti, Mauro Gianni Perrucci

## Abstract

**Objective:** Bilingualism has been associated with cognitive benefits and a potential protective effect against neurodegenerative conditions. Previous research has shown that bilingual individuals exhibit greater white matter integrity compared to monolinguals of the same age. However, the impact of foreign-language learning on brain structure in older adults during the initial stages of language acquisition remains uncertain. This study aimed to investigate the cognitive and structural effects of a four-month-long foreign language learning program in a group of healthy monolinguals older adult.

**Materials and Methods:** Thirteen Italian-speaking participants (aged 59-78) underwent a four-month intensive English course for beginners. Pre- and post-assessments were conducted to evaluate executive cognitive functions, as well as diffusion tensor imaging (DTI) to examine brain structural changes.

**Results:** The study findings showed substantial increases in axial, radial, and mean diffusivity during the four-month language learning period. The most prominent variations were observed in key brain regions, namely the fronto-occipital fasciculus, the superior longitudinal fasciculus, and the corpus callosum areas. Notably, brain-behaviour correlations indicated a robust positive relationship between changes in axial diffusivity and performance on the Stroop task, a well-established measure of cognitive interference inhibition.

**Conclusions:** These findings suggest that a four-month foreign language learning program can lead to structural changes in the brain, particularly affecting white matter integrity and that these structural changes are associated with improvements in executive functions. The study underscores the potential of brief language learning interventions to influence brain structure and enhance cognitive abilities in older adults.

**Highlights:**

- Bilingualism is associated with cognitive advantages and the potential for protection against neurodegenerative conditions, as evidenced by bilingual individuals displaying enhanced white matter integrity compared to individuals who are monolingual and of the same age.
- The study investigates how a four-month foreign language learning program affects brain structure in older adults during the initial stages of language acquisition
- The research reveals significant increases in axial, radial, and mean diffusivity in key brain regions, including the fronto-occipital fasciculus, superior longitudinal fasciculus, and corpus callosum, during the language learning period.
- The study demonstrates a positive link between changes in axial diffusivity and performance on the Stroop task, indicating that short language learning interventions can lead to structural brain changes associated with improved executive functions in older adults.

## Introduction

The aging population is experiencing rapid growth, and advancements in healthcare have led to significant improvements in life expectancy. However, cognitive decline and the development of neurodegenerative conditions, such as Alzheimer’s disease, Parkinson’s disease, and various forms of dementia, are primarily associated with the aging process. These conditions profoundly impact the functional integrity and connectivity of the brain, necessitating the identification of both pharmacological and non-pharmacological interventions to mitigate age-related brain changes and delay the onset of neurodegenerative disorders (Buckner, 2004; Herrup, 2010).

Pathological aging and the emergence of dementia significantly disrupt the functional and structural connectivity processes of the brain.

Consequently, a pivotal objective of modern medicine is to identify potential interventions, both pharmacological and non-pharmacological, that can counteract age-related brain changes and delay the onset and progression of neurodegenerative conditions (Granzotto and Zatta, 2014; Prakash et al., 2015; Pruchno and Carr, 2017).

By focusing on these underlying mechanisms, interventions aim to preserve cognitive function and enhance overall brain health in older adults. Recent scientific and technological advances have revealed that age-related dysfunctional brain processes can be slowed due to the remarkable ability of the nervous system to adapt and reorganize its structure, functions, and connections in response to intrinsic and extrinsic stimuli, a phenomenon known as neural plasticity (Hertzog et al., 2008; Hillman et al., 2008; Mora, 2013; Phillips, 2017).

This remarkable capacity for neural plasticity opens new possibilities for interventions aimed at mitigating age-related brain changes. Moreover, emerging evidence suggests that certain lifestyle factors, such as higher levels of education, occupational attainment, regular exercise, and sustained engagement in cognitively challenging activities, play a critical role in slowing down the process of cognitive aging, which is associated with the decline in cognitive abilities (Baltes et al., 2006; Boucart et al., 2015;Erickson et al., 2011; Foubert-Samier et al., 2012; Hultsch et al., 2008; Pieramico et al., 2014;Singh-Manoux et al., 2012).

The prevailing hypothesis in the field proposes that interventions aimed at mitigating age-related cognitive decline primarily exert their effects by modulating the brain and enhancing cognitive reserve. (Krivanel et al., 2021; Stern, 2021; Perneczky, 2022). Cognitive reserve, a protective mechanism, is considered a deposit account of sorts, linking engagement in cognitively challenging tasks to better cognitive aging at both the physical brain level (brain reserve) and cognitive performance (cognitive reserve) (Stern, 2002; Stern, 2021). It involves efficient connectivity among neural circuits, supporting sustained cognitive functions even in the face of age-related decline, injury, disease, or their combination. Cognitive reserve provides behavioural compensation, enabling greater cognitive performance than expected given one’s aging or degraded brain (Stern, 2021). Experimental evidence has demonstrated the effectiveness of non-pharmacological approaches, such as physical exercise or cognitive stimulation, in promoting the maintenance of brain and cognitive reserves (Brem and Sensi, 2018; Ngandu et al., 2015; Vecchio et al., 2018). However, the brain’s functional and structural changes associated with these neuroprotective effects remain poorly understood (Alwin and Hofer, 2011; Barulli and Stern, 2013; Campos-Esparza et al., 2009; Gelfo et al., 2018; Sensi et al., 2018; Sisalli et al., 2015).

Bilingualism or multilingualism has also emerged as a significant factor in preserving brain integrity against age-related changes (Abutalebi et al., 2014a, b; Anderson et al., 2018; Gallo et al., 2022; Gallo and Abutalebi, 2023; Voits et al., 2022; Schönpflug, 2001). Bilingualism exhibits critical neuroprotective properties and plays a significant role in modulating executive functioning (Gallo et al., 2020). Recent findings indicate that the ability to speak a second language from birth or learn a second language during development provides a high degree of neuroprotection later in life (Del Maschio et al., 2019; Sulpizio et al., 2020). Differences are observed in terms of brain functional and structural changes, as well as the maintenance and enhancement of cognitive abilities (Gabryś-Barker, 2017; Pot et al., 2019a; Schlegel et al., 2012; Valis et al., 2019; Wong et al., 2019). In particular, it has been demonstrated that learning a foreign language in adulthood can improve global cognition and functional connectivity (Antoniou et al., 2013; Bak et al., 2014; Bubbico et al., 2019; Nijmeijer et al., 2021, Klimova and Pikhart, 2020; Pliatsikas et al., Schweizer et al., 2012; Ware et al., 2017). Moreover, studies conducted by Keijtzer and colleagues explored language learning as a potential tool against cognitive impairment, focusing on socio-affective factors and positive effects in Late-Life Depression (Pot et al., 2019b, 2019c).

Importantly, bilingualism has shown potential in delaying the manifestation and diagnosis of Alzheimer’s disease. Meta-analysis studies have indicated that age-related cognitive decline is delayed by approximately 4.5 years in bilingual individuals compared to age-matched monolinguals (Bialystok et al., 2007; Calabria et al., 2020; Craik et al., 2010; Brini et al., 2020; Paulavicius et al., 2020). A large meta-analysis by Anderson et al. (2020) demonstrates that bilingualism is protective in delaying symptoms regardless of educational level and socioeconomic status.

Despite the plethora of evidence suggesting that bilingualism is protective against cognitive decline in older age, there is limited knowledge regarding the specific neural changes resulting from foreign-language learning that led to these protective outcomes. Furthermore, there is a significant gap in longitudinal studies examining the effects of foreign language learning on structural brain changes and cognition in healthy older adults. It remains unclear whether foreign-language learning can provide neuroprotection when administered later in life (Freedman et al., 2014; Perani and Abutalebi, 2015; Pliatsikas et al., 2020).

To accurately assess the impact of interventions on brain neuroplasticity, it is essential to employ a quantitative technique that can effectively detect subtle changes in brain structure. Existing research provides compelling evidence indicating a correlation between brain aging and alterations in the microstructure of white matter (WM), which could potentially contribute to cognitive decline. WM tracts, coherent bundles of myelinated fibers, serve as the structural foundation for neural communication, facilitating the transmission of information between different brain regions. Magnetic Resonance Imaging (MRI) has been extensively employed to assess and investigate brain and WM structural changes underlying age-related processes, (Cole and Franke, 2017; Gracien et al., 2017; Ward et al., 2015). In particular, the microstructural integrity of WM has been assessed by using Diffusion tensor imaging (DTI), a non-invasive and quantitative method exploiting the Brownian motion of water molecules in brain tissues (Assaf and Pasternak, 2008; Borrelli et al., 2019). DTI measures (mean diffusivity, axial/radial diffusivity, fractional anisotropy) provide information about the integrity and organization of WM tracts. WM lesions have been implicated in cortical structural impairment and the deterioration of neuronal connections (Bartzokis et al., 2004; Cox et al., 2016; Le Bihan, 2014; O’Sullivan et al., 2001). The differences in brain WM structure in terms of DTI-derived quantitative parameters can be quantified by using the Tract-Based Spatial

Statistics (TBSS) method (Marrale et al., 2016), which allows to perform voxel-wise analysis of diffusion data in multiple subjects. Moreover, correlating DTI parameters with cognitive parameters allows us to understand how changes in WM integrity may be associated with variations in cognitive performance, thus elucidating the relationship between brain structure and cognitive function, and providing insights into the neural underpinnings of cognitive processes and how they are shaped by brain connectivity.

In a previous study conducted, a cohort of older adults participated in a 4-month foreign language learning course (beginner-level English course, 2 hours/week). The study demonstrated that foreign-language learners exhibited improved performance in global cognitive functions as well as functional connectivity in the language and executive networks compared to the control group (Bubbico et al., 2019). Specifically, enhancements were observed in the right inferior frontal gyrus (rIFG), right superior frontal gyrus (rSFG), and left superior parietal lobule (lSPL).

In this study, the aim was to build upon these previous findings by investigating white matter (WM) structural modifications using Diffusion Tensor Imaging (DTI). Additionally, drawing from numerous studies conducted on bilingual, trilingual, and multilingual subjects (Antón et al., 2019; Filippi et al., 2012; Poarch and van Hell, 2012), a focus was placed on cognitive tests designed to assess executive functions, such as the Stroop task. It is well-documented that executive functions receive extensive training in individuals proficient in two or more languages. Bilinguals consistently demonstrated superior performance compared to monolinguals, despite similar outcomes on other neuropsychological measures (Bialystok and Poarch, 2014; Coderre and van Heuven, 2014; Hilchey and Klein, 2011). Examining changes in WM tracts connecting gray matter (GM) regions can provide valuable insights into the connectivity changes and neuroprotection observed in the cohort of foreign-language learners. Moreover, we can gain a better understanding of how language training impacts the structural connectivity of the brain and how these changes may contribute to the observed functional connectivity changes. Furthermore, changes in white matter integrity have been linked to cognitive functioning and the capacity to resist age-related cognitive decline. Therefore, studying WM changes can offer important complementary information to further elucidate the findings observed in our cohort of foreign-language learners. The examination of WM tracts can provide insights into potential mechanisms of neuroprotection and help establish a more comprehensive understanding of the neural adaptations occurring as a result of foreign-language learning.

## Materials and methods

### Study participants

Fifteen monolingual Italian speakers, with a mean age of 69 years (standard deviation, SD 5.6 years), were recruited and selected through advertisements for participation in the study (**Table 1**). The enrolled participants consisted of both genders and were right-handed individuals. The educational level was recorded in terms of years of education for each participant. Prior to inclusion, all participants underwent a thorough screening interview, which included medical and neuropsychological assessments. Exclusion criteria comprised suspected cognitive decline based on the neuropsychological assessment, presence of conditions that could impede safe engagement in the intervention (such as depression, neurodegenerative or psychiatric disorders, symptomatic cardiovascular disease, severe self-reported sensory impairments, ongoing participation in another intervention trial), and any contraindication to MRI scanning (including metal implants and claustrophobia). Individuals who were smokers, exhibited behavioral or substance abuse issues, or reported recent consumption of caffeine or alcohol within 24 hours prior to the MRI session were also excluded from control for external factors. The included participants underwent two brain MRI scans, before and after a 4-month period of foreign language learning (see below for further details). Informed consent was obtained from all participants, and the study was approved by the Institutional and Ethical Committee of the University “G. d’Annunzio” of Chieti-Pescara. Detailed demographic characteristics of the participant cohort can be found in **Table 1**.

**Table 1.** Baseline characteristics of the study participants.

|  | Mean (SD) |
| --- | --- |
| **Participants (n)** | 13 |
| **Age (years)** | 69 (5.6) |
| **Sex (M/F)** | 2/11 (Y.=14.3) |
| **Education (years)** | 9.4 (2.7) |
| **MMSE (score)** | 27.17 (1.71) |

SD = standard deviation; MMSE = Mini-Mental State Examination test score

### Study design

The foreign language learning course was conducted by a qualified native English teacher. The program followed a structured format, consisting of once-a-week group sessions over a period of 4 months, totaling 16 weeks. Each class was approximately 1.5 hours long, including a 15-minute break, and was supplemented with 30 minutes of homework exercises for further practice. The program was built upon a protocol that had already proven functional changes in our previous study (Bubbico et al., 2019).

The course curriculum focused on various aspects of language acquisition, including building basic vocabulary, grammar, speaking, and writing skills. Additionally, participants were introduced to British and American English traditions and culture. Lessons were designed as collaborative projects, offering ample opportunities for oral and written communication practice. The participants specifically worked on developing their grammar and vocabulary in areas such as culture, travel, shopping, and family. To monitor progress, qualitative assessments were conducted by the native teacher at the beginning and end of the course.

Throughout the 4-month training session, participants were instructed to maintain their regular daily routines. Within two weeks before and after completing the foreign language course, all subjects underwent cognitive evaluations and MRI scanning to assess potential changes in cognitive functioning and brain structure.

### Neuropsychological and behavioral assessment and analysis

All participants underwent executive function neuropsychological testing using the Stroop Colour and Word Test (SCWT) both before and after the training period. The Stroop test is a well-established measure of executive abilities (Caffarra et al., 1991; Scarpina and Tagini, 2017; Treisman and Fearnley, 1968).

In particular, the SCWT is a widely used neuropsychological test that assesses the ability to inhibit cognitive interference, known as *The Stroop Effect* (Brugnolo et al., 2016; Scarpina and Tagini, 2017). In this study, the original paper printed version of the ninety-item paradigm with oral responses was employed, following the methodology described by Stroop (1935) and Penner et al. (2012). The test consisted of three conditions: the “reading condition,” the “denomination condition,” and the “interference condition.” Each condition comprised three pages with ten stimuli per page.

In the “congruous condition,” participants were required to read color-words (green, blue, and red) printed in black ink (W) as quickly as possible. Then, they were asked to name different color patches (C). In contrast, the “color-word (CW) condition” presented color-words printed in an inconsistent color ink (e.g., the word “red” printed in green ink). In this incongruent condition, participants had to name the color of the ink instead of reading the word, thus inhibiting interference from the more automated task of reading (MacLeod and Dunbar, 1988; Ivnik et al., 1996). This interference inhibition difficulty is known as the Stroop effect (Stroop, 1935). Although the SCWT primarily measures the ability to inhibit cognitive interference, previous research has also applied it to assess attention, processing speed, cognitive flexibility (Jensen and Rohwer, 1966), and working memory (Kane and Engle, 2003), indicating its potential for measuring multiple cognitive functions. Participants were instructed to respond as quickly and accurately as possible, with the experimenter recording reaction times (RT) using a stopwatch. The variables of interest were the reaction times and the number of errors made in the three test conditions.

Descriptive statistics, including arithmetic mean, standard deviation, median, and percentage, were used to report the general characteristics of the study population (**Table 1**). Pre- and post-intervention comparisons of the neuropsychological scores were conducted using the general linear model. The analyzed outcomes were reported as the difference between the pre- and post-intervention scores. Statistical significance was determined by a two-tailed P-value p < 0.05, and Bonferroni’s correction was applied to adjust the significance threshold for multiple comparisons. Data analysis was performed using Statistica 12.0.

### Imaging procedure

The study utilized structural MRI data collected with a Philips Achieva 3 Tesla scanner (Philips Medical Systems, Best, Netherlands). The MRI scans were conducted using a 3D fast field echo T1-weighted sequence to acquire a high-resolution structural volume. The parameters for the T1-weighted sequence were as follows: sagittal acquisition, matrix size of 240 × 240, field of view (FOV) of 256 mm^2^, slice thickness of 1 mm without any gap, in-plane voxel size of 1 × 1 mm^2^, flip angle of 8°, repetition time (TR) of 8.2 ms, and echo time (TE) of 4 ms. The imaging parameters for diffusion-weighted imaging (DWI) were as follows: TR/TE = 6382 ms/70 ms, FOV = 224 × 120 × 224 mm³ covering 60 oblique axial slices without any gap, voxel size of 2 × 2 × 2 mm³, and total scan duration of 494 s, b=0,1500 s/mm^2^, 60 non collinear diffusion directions, 1 image without diffusion weighted (b0). The MRI acquisitions (structural and DWI) were performed at baseline and after the four-month language course.

### DTI and Tract-Based-Spatial-Statistics (TBSS) analysis

The DICOM conversion to NIfTI format was performed using the dcm2nii tool from MRIcron (Jenkinson et al., 2012; Smith et al., 2004). Image denoising was performed in MRtrix3 (Tournier et al., 2019), followed by eddy-current induced distortion correction and motion correction in FSL (FMRIB’s FSL 5 toolbox, Woolrich et al., 2009). The gradient table was corrected after eddy-current correction. Brain extraction was performed using FSL’s BET function, employing the corrected b0 image and applying this extraction to the rest of the diffusion-weighted volumes., The denoised, corrected and skull-stripped diffusion data were used to derive the diffusion tensor and generate maps for fractional anisotropy (FA), axial diffusivity (AD), mean diffusivity (MD), and radial diffusivity (RD) (Basser et al., 1994, Behrens et al., 2003).

The diffusion parametric maps (FA, MD, AD, RD) were then fed into the Tract-Based Spatial Statistics (TBSS) toolbox, to perform voxel-wise analysis with a sample size of n = 50000 and an FA threshold of 0.2. TBSS aligned individual subjects’ FA maps to a common WM skeleton, allowing for voxel-wise comparison of diffusion measures (MD, AD, RD, FA) along the skeleton, to verify whether the intervention (language training course) induced any changes in the WM tracts or regions. Moreover, the parametric maps are projected onto a standardized space (Montreal Neurological Institute, MNI space). FreeSurfer was used for segmentation of brain into three tissues (WM, gray matter, GM, and cerebro-spinal fluid), using the structural image. The GM and WM were further parcellated. The parcellated atlases obtained were then projected onto the diffusion native space and onto the standard MNI space, using FSL FLIRT. TBSS was used to identify significant clusters (pre-vs post-treatment) in the DTI parameters in a whole brain analysis. Subsequently, those clusters were used as regions of interest (ROIs).

### Association between TBSS results and cognition

A Pearson’s correlation analysis was conducted to explore the association between executive abilities and white matter microstructure. Executive abilities were assessed using Stroop scores corrected for age and education, including reaction times (RT) and the number of errors. The DTI parameters in white matter (AD, RD, MD, and FA) were utilized to measure the diffusion of water molecules in brain tissues, providing crucial insights into white matter tract integrity and organization.

By correlating DTI parameters with cognitive measures, a relationship between brain structure and cognitive function was investigated. This approach allows for understanding how changes in white matter integrity, as reflected by DTI measures, may be associated with variations in cognitive performance. Such correlations offer valuable insights into the neural underpinnings of cognitive processes and their modulation by brain connectivity.

The analysis accounted for covariates, including sex, age, and educational level. The statistical analysis was performed using the Statistical Package for Social Sciences (SPSS, Inc., Chicago) version 15.0.T. A significance level of p < 0.05 was set for the correlation analysis, and Bonferroni correction was applied to account for multiple comparisons.

## Results

### Stroop test performance and DTI metrics

Participants underwent pre (T0) and post (T1) testing using the Stroop Test.

The results indicated that there were no significant improvements in error rates or reaction times (RT) from T0 to T1 (both p = 0.24, **Fig. 1**).

**Figure 1.**
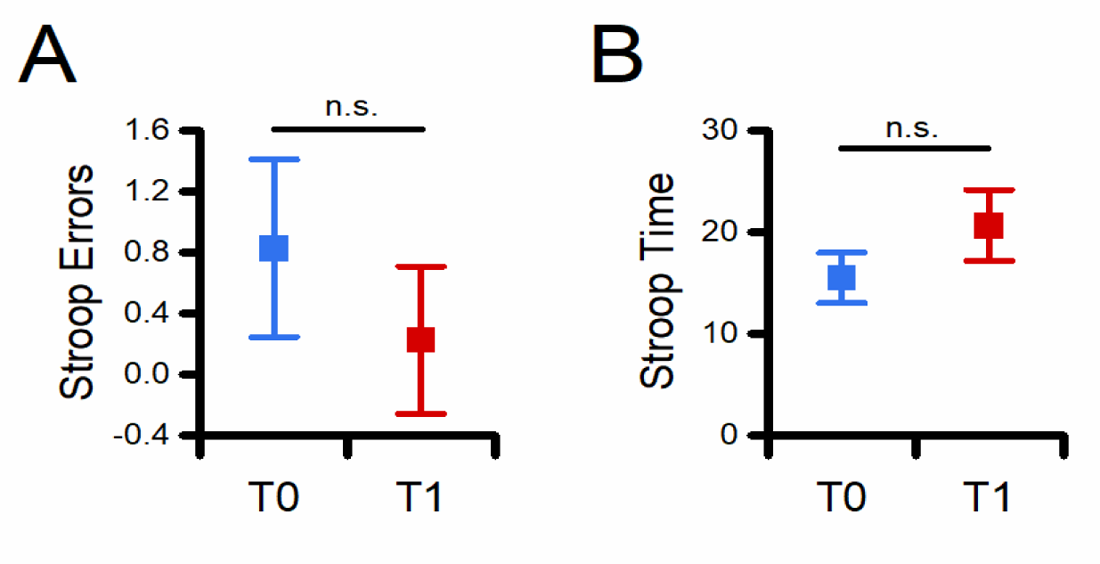
Foreign language learning and cognitive flexibility results. The graphs depict changes in Stroop test errors (A) and reaction time (B) before and after the language training course (T0 and T1, respectively). Data are represented as mean ± standard error of the mean (SEM). n.s. = not significant.

For the analysis of WM integrity, the findings revealed a statistically significant increase in the values of AD and MD after the 4-month language training, while no significant change was observed for RD and FA (**Fig. 2**). Regions of significant changes in DTI parameters were identified in fronto-occipital fasciculus, the superior longitudinal fasciculus, and the corpus callosum areas. The coordinates of such regions in the Montreal Neurological Institute (MNI) space are listed in **Table 2**. These coordinates represent the brain locations where significant pre-post language training changes were observed for AD and MD.

**Figure 2.**
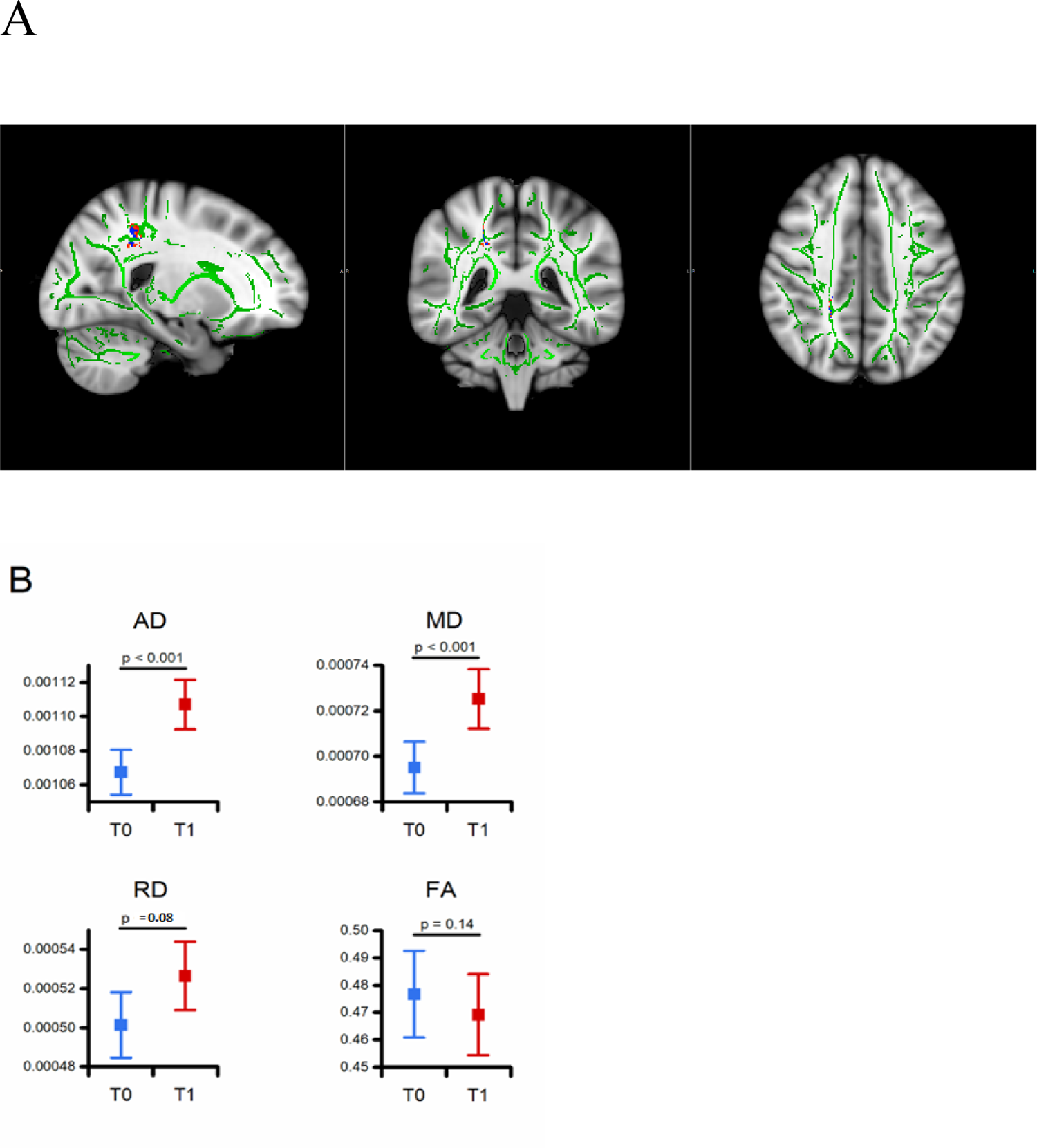
DTI parameters before and after the language training. (A) The pictogram illustrates the sagittal (left), coronal (middle), and axial (right) views of the TBSS analysis. Green depicts the FA skeleton, common to all the subjects. Red and blue depict the AD and MD significative areas, respectively. (B) Graphs depict quantification values for AD, MD, RD, and FA, as indicated, before (T0) and after (T1) the training course. Data are represented as mean ± SEM. R, right; L, Left.

**Table 2.** Coordinates in the Montreal Neurological Institute (MNI) space for areas with significative pre-post changes in DTI-parameters.

| Coordinates | MNI |
| --- | --- |
| **AD** | 22 -43 45 |
| **MD** | 20 -42 38 |

AD= axial diffusivity, MD= mean diffusivity; MNI= Montreal Neurological Institute.

To gain insights into the patterns of associations between variables, helping to identify potential relationships and further understand the underlying processes or mechanisms at play, correlation matrix was conducted to explore associations between changes in the DTI parameters from T0 to T1. Significant positive correlations were found between changes in AD and MD (r = 0.80, p < 0.001, Table 3), as well as between RD and MD (r = 0.90, p < 0.001). Additionally, a negative correlation was observed between RD and FA (r = −0.60, p = 0.03).

**Table 3.**
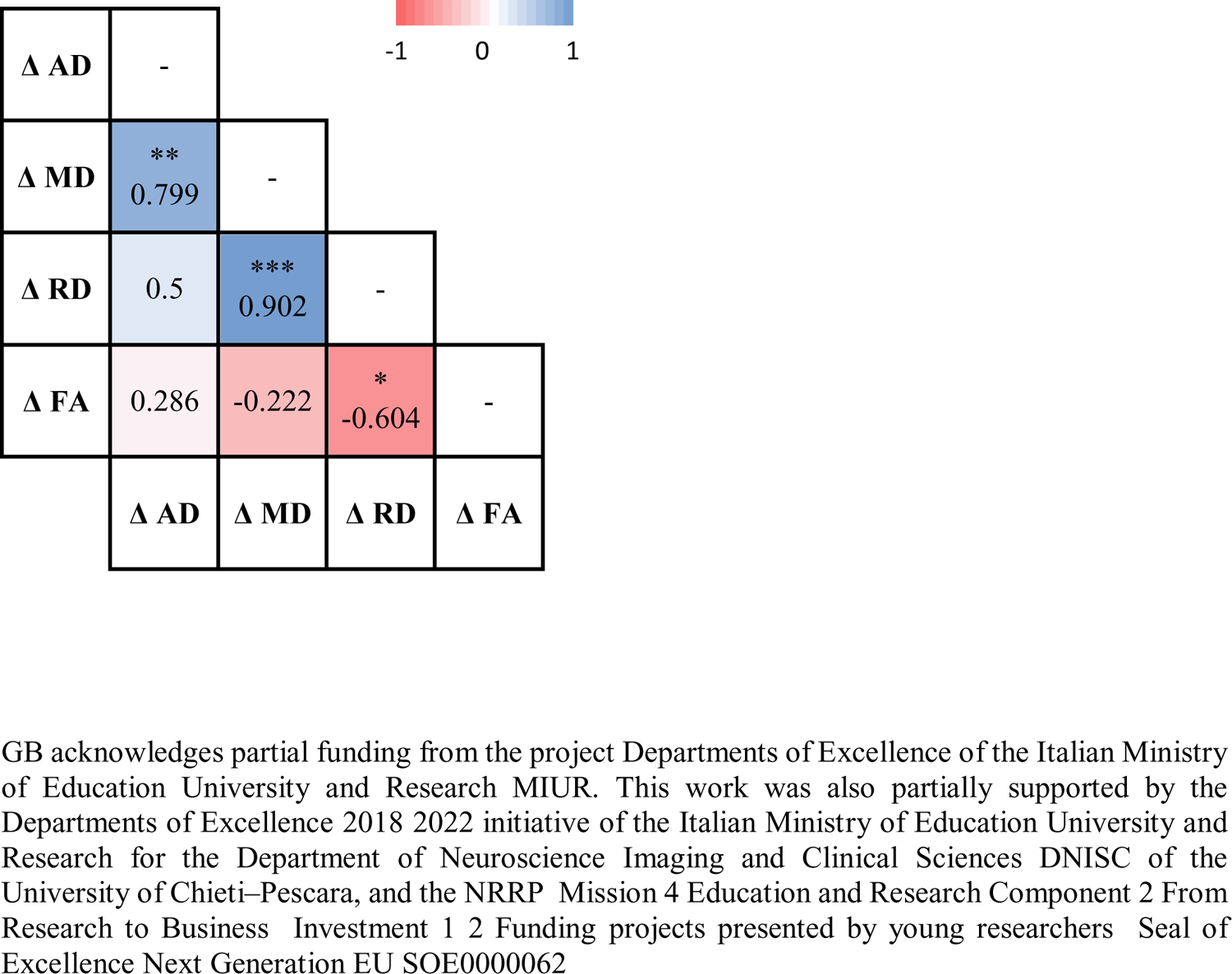
Correlation matrix of the differences between T0 and T1 of AD, MD, RD, and FA. * Indicates p < 0.05, ** indicates p < 0.01, and *** indicates p < 0.001.

|  |  |  |  |  |
| --- | --- | --- | --- | --- |
| $\Delta$ AD | - | | | |
| $\Delta$ MD | **<br>0.799 | - | | |
| $\Delta$ RD | 0.5 | ***<br>0.902 | - | |
| $\Delta$ FA | 0.286 | -0.222 | *<br>-0.604 | - |
| | $\Delta$ AD | $\Delta$ MD | $\Delta$ RD | $\Delta$ FA |

### Association between Stroop test and white matter integrity

Although there was no significant change in overall response times on the Stroop task from T0 to T1, an interesting finding emerged regarding the association between changes in Stroop task performance and changes in axial diffusivity (AD). Specifically, a strong negative correlation was observed between the change in Stroop task performance and the change in AD (r = −0.67, p = 0.01). This implies that participants who exhibited an increase in AD throughout the training period also demonstrated faster response times on the Stroop task. However, no such relationship was found between AD and error rates on the Stroop task. Additionally, no significant relationships were observed between the Stroop task and the other DTI metrics.

## Discussion

This study aimed to examine the impact of foreign language learning on WM integrity in a small sample of healthy older adults. The investigation disclosed three key findings. Firstly, foreign language learning resulted in increased axial diffusivity and mean diffusivity, indicating positive changes in white matter structural integrity. Secondly, changes in axial diffusivity and radial diffusivity over the course of training were positively correlated with changes in mean diffusivity, while changes in radial diffusivity were negatively correlated with changes in fractional anisotropy. Lastly, improvements in Stroop task performance were associated with changes in axial diffusivity.

Results indicate that during the four-month language learning period, there were significant changes in diffusivity measures within the brain, particularly in fronto-occipital fasciculus, the superior longitudinal fasciculus, and the corpus callosum areas. Axial diffusivity refers to the diffusion of water molecules along the axonal fibers of the brain, while radial diffusivity refers to diffusion perpendicular to the axonal fibers. Mean diffusivity is a measure of overall diffusion in multiple directions. The significant increases in axial, and mean diffusivity suggest changes in the microstructure of the brain’s white matter, which may reflect increased axonal branching, myelination, or other structural modifications.

Learning new skills, including foreign languages, can lead to neuroplasticity, the brain’s ability to reorganize itself by forming new connections and modifying existing ones. As a result of foreign language learning, the brain may have strengthened existing white matter connections or formed new ones, leading to the observed increases in axial diffusivity and mean diffusivity.

The fronto-occipital fasciculus (FOF), the superior longitudinal fasciculus, and the corpus callosum areas seem to be particularly affected by these changes. The FOF runs longitudinally, connecting the frontal cortex, which is responsible for higher cognitive functions such as decision-making, planning, and social behaviour, with the occipital cortex, which plays a crucial role in visual processing and perception. Moreover, it is involved in several functions, including visual processing, executive functions, language processing, and spatial cognition (Friedman and Robbins., 2021).

The FOF plays a critical role in transmitting visual information from the occipital lobe, where visual stimuli are initially processed, to the frontal lobe. This pathway is involved in the integration and interpretation of visual information, allowing to recognize and make sense of the world. The frontal lobe is heavily involved in executive functions, higher-order cognitive processes that regulate and control other cognitive abilities. These functions include decision-making, problem-solving, working memory, attention, and cognitive flexibility. The FOF’s connections with the frontal cortex are important for coordinating and facilitating these executive functions. Some studies suggest that the FOF may also play a role in language processing, particularly in the integration of visual and linguistic information during reading and comprehension tasks (Dato et al., 2023). However, the exact role of the FOF in language processing is still a topic of ongoing research and debate. Spatial cognition involves the perception, representation, and processing of spatial information. The FOF’s connections with various brain regions, including those involved in spatial processing, may contribute to our ability to navigate and understand spatial relationships.

The superior longitudinal fasciculus (SLF) connects regions in the two hemispheres of the brain. This interhemispheric connectivity allows for information transfer and integration between homologous regions on the left and right sides of the brain. The SLF plays a crucial role in integrating sensory information with motor functions. It helps in coordinating sensory perception and motor responses, enabling interaction with the environment. Components of the SLF have been implicated in language-related functions. Specifically, SLF is thought to be involved in aspects of language comprehension and semantic processing. SLF has also been associated with visuospatial processing and attention. This pathway helps in perceiving and understanding spatial relationships, which are essential for tasks such as navigation, object recognition, and mental imagery (De Benedictis et al., 2016). Some portions of the SLF are linked to working memory and executive functions. Working memory is the ability to temporarily hold and manipulate information for cognitive tasks, while executive functions involve higher-order cognitive processes like decision-making, planning, and cognitive flexibility. Moreover, studies show that SLF is also believed to be involved in attentional processes and cognitive control, allowing individuals to focus their attention, suppress irrelevant information, and switch between different tasks or mental states. Overall, the SLF is a crucial white matter pathway that plays a central role in facilitating communication and coordination between various cortical regions, contributing to a wide range of cognitive functions and behaviors.

The corpus callosum plays a crucial role in facilitating communication between the two hemispheres. It allows both hemispheres to share information, process sensory inputs, and coordinate motor outputs. This communication is essential for integrated brain function, as it allows the two hemispheres to work together as a unified system. Information processing in the brain often involves multiple regions in both hemispheres. The corpus callosum facilitates the integration of cognitive functions, such as language processing, visuospatial skills, memory, and attention, by allowing relevant information from one hemisphere to be shared with the other.

Some studies suggest that the corpus callosum may also play a role in emotional processing and empathy (Anderson et al., 2017). While language processing involves multiple brain regions, the corpus callosum is particularly important for certain aspects of language function, such as the transfer of linguistic information between the left hemisphere (often dominant for language in most individuals) and the right hemisphere. Results support the involvement of these areas of the white matter in our training.

The study observed a significant improvement in both the Stroop test error rate and reaction times after the training period. The Stroop test is a well-known psychological task that assesses cognitive control and executive functions. The significant improvement in the Stroop test performance suggests that the training had a positive impact on cognitive control and executive functions. Learning and practicing new skills, such as the cognitive tasks involved in the training, can lead to neuroplasticity. Neuroplasticity refers to the brain’s ability to reorganize and adapt by forming new connections between neurons. As participants engaged in the training, it is likely that their brain circuits associated with cognitive control tasks were strengthened and refined, resulting in improved performance on the Stroop test. The training might have enhanced participants’ attentional skills and working memory capacity. Attention is crucial for filtering out distractions and maintaining focus on relevant information while working memory enables the temporary storage and manipulation of information during complex tasks like the Stroop test. Improved attention and working memory capacity would have contributed to faster and more accurate responses on the Stroop test.

Regarding the changes in axial diffusivity (AD) being associated with changes in Stroop test performance, it suggests a potential relationship between white matter integrity and cognitive performance. Changes in AD are indicative of alterations in the diffusion of water molecules along the main axis of white matter fibers.

However, it’s crucial to acknowledge that the precise mechanisms and interplay among white matter integrity, cognitive performance, and training effects can be intricate and multifaceted, requiring further investigation. Additional research is needed to further investigate the underlying neural mechanisms and to replicate and validate these findings in larger and more diverse samples.

In conclusion, although not significant, the observed trends of the improvement in the Stroop test performance after training could be attributed to neuroplasticity, improved attention and working memory, and task-specific learning. Instead, significant changes in AD and Stroop test performance highlight the potential link between white matter integrity and cognitive functions, which warrants further investigation.

The lack of reaching statistical significance despite the observed improvements in the Stroop test performance could have been attributed to several factors. The number of participants involved in the research could have influenced the ability to detect statistically significant effects. Individual differences in cognitive abilities, neural architecture, and response to training might have introduced variability in the data, potentially masking the true effect of the training on cognitive performance. Future studies that take into consideration these points are needed. The sensitivity of the measurement methods used in the study could have affected the ability to detect significant changes and might have had limitations in detecting subtle changes in cognitive performance. The duration of the training period might have also played a role in the lack of significance. The training was relatively short, and the effects on cognitive performance might not have reached a level of statistical significance within that timeframe.

Furthermore, the study found a strong positive association between changes in axial diffusivity and performance on the Stroop task. The Stroop task measures cognitive interference inhibition, which refers to the ability to suppress irrelevant or conflicting information. The positive correlation suggests that the structural changes in the brain, as reflected by increased axial diffusivity, may be related to improved cognitive performance on the Stroop task.

In summary, these findings suggest that foreign language learning can lead to significant changes in brain microstructure, particularly in the fronto-occipital fasciculus, the superior longitudinal fasciculus, and the corpus callosum areas. The observed associations with cognitive performance indicate that these structural changes may have functional implications for cognitive abilities such as inhibitory control. These results suggest that a 4-month foreign language course exposure can enhance executive functions by promoting positive changes in white matter integrity.

Our findings are consistent with and build upon our previous research, which demonstrated brain functional changes measured by fMRI (Bubbico et al, 2019). Taken together, these results highlight foreign language learning as a powerful facilitator of neuroplasticity-related changes in the aging brain. Aging and the associated cognitive decline are widely recognized as primary risk factors for dementia (Herrup, 2010). Considering the lack of effective pharmacological interventions to delay the progression of neurodegenerative conditions, non-pharmacological approaches, such as cognitive training, can be employed as cost-effective and efficient strategies. (Kivipelto et al., 2018; Joshi et al., 2022; Miao et al., 2022; Winblad et al., 2016; Palmer et al., 2022). This approach is based on the concept of the brain’s “reserve,” which encompasses cognitive and neurobiological processes that promote resilience and maintain optimal brain function (Cabeza et al., 2018). Notably, various factors, including lifelong education, adaptive experiences, lifestyle, and intelligence, can modulate this reserve during aging (Gallo et al., 2020; Ory et al., 2003; Park and Gutchess, 2002; Reuter-Lorenz and Lustig, 2005; Stern, 2002). Hence, the stimulation of this cognitive reserve may result in enduring brain alterations that help maintain cognitive function in older adults. A growing body of research is investigating the optimal combination of cognitive tasks to achieve the most favorable outcomes. Our study provides support for the brain reserve hypothesis and the positive impact of foreign language learning on cognitive fitness (Alladi et al., 2013; Bak and Alladi, 2014; Bialystok et al., 2012; Luk et al., 2011). Specifically, our foreign language learning protocol resulted in increased axial diffusivity (AD), and greater increases in AD were associated with improved performance on the Stroop task in terms of response times. AD reflects diffusion along the fiber direction in white matter and is linked to favorable outcomes. These findings are consistent with a recent study that demonstrated bilingual individuals have higher AD values compared to monolingual individuals after controlling for potential confounding factors (Anderson et al., 2018a).

In a quantitative meta-analysis of functional neuroimaging studies, Decety and colleagues (2007) found that the right inferior parietal cortex is involved in lower-level computational processes related to the sense of agency and attentional reorientation to salient stimuli. Considering that bilingualism enhances performance on attentional tasks (Bialystok, 2017, Grundy, Anderson, Bialystok, 2017), microstructural changes in this white matter tract may serve as a contributing mechanism to this enhancement. Furthermore, our study revealed an increase in radial diffusivity (RD) over time, which parallels the increase in axial diffusivity (AD) and provides an explanation for the lack of changes in fractional anisotropy (FA) values.

In contrast to the findings of Rossi et al. (2017), although in different areas than those observed in our study, the lack of significance in FA could be attributed to the relatively small sample size and the short duration of the language course. Furthermore, the more comprehensive and diverse curriculum of our course, which extended beyond the mere acquisition of a hundred words, could have imposed a heightened cognitive demand for processing and memorization.

AD and RD represent opposing outcomes, with AD reflecting microstructural integrity and higher RD values indicating white matter damage. It is possible that some individuals in our study were experiencing age-related physiological decline during the 4-month period, as indicated by changes in RD. The increase in AD may have occurred as a result of foreign language learning and compensated for any increase in RD. The positive association between AD and Stroop task performance over time, despite no significant change in overall group performance, supports this interpretation. This idea is further supported by our previous work, which demonstrated significant improvement in functional connectivity in the same foreign-language learning group compared to matched controls from the same population (Bubbico et al., 2019). These findings suggest that the increase in AD may be an underlying mechanism that contributes to the advantage of bilinguals over monolinguals in executive function tasks more frequently than expected by chance (meta-analysis in Grundy, 2020). Additionally, more efficient communication between gray matter regions through white matter tracts may contribute to cognitive reserve in older age and potentially delay cognitive decline.

Compelling evidence indicates that early bilinguals outperform monolinguals on various executive function tasks, particularly those related to inhibitory functions (e.g., Souza and Souza, 2016; Green and Abutalebi, 2013). Our findings extend this view by demonstrating that neuroplasticity and associated changes in executive functions can also be observed in older individuals learning a foreign language later in life. Participants who exhibited the greatest increases in AD from the beginning to the end of the foreign-language learning course also showed the most significant improvements in response speed on an inhibitory control task (i.e., Stroop task). It is possible that the observed structural changes were not sufficient to trigger detectable improvements in executive functions. However, given the relatively short duration of the intervention, it is likely that our intervention stimulated subtle plasticity-related mechanisms that did not result in detectable changes in executive functions. Perhaps with a longer learning process, these results could have been enhanced. Another hypothesis is that the technical limitations of the tests employed in our study may have masked more nuanced changes in executive functions, and a more comprehensive and detailed battery of tests could have revealed enhanced changes.

Importantly, the loss of executive functions is an early event in the processes leading to pathological brain aging (Buckner, 2004b; Murman, 2015). Individuals with impaired executive performance are more likely to develop conditions associated with dementia (Harciarek et al., 2017; Ravdin et al., 2013; Perry and Hodges, 1999; Tarawneh and Holtzman, 2012).

In a previous study conducted by our group, we demonstrated functional brain changes in the right inferior frontal gyrus (rIFG), right superior frontal gyrus (rSFG), and left superior parietal lobule (lSPL) following foreign language learning (Bubbico et al., 2019). However, functional changes alone may be transient without robust and enduring structural modifications (Herold et al., 2019; Lisman et al., 2018; Oberman & Pascual-Leone, 2013). To investigate the effect of training at the structural level, we employed diffusion tensor imaging (DTI), which allows for the examination of white matter (WM) microstructural changes. The rationale for assessing WM changes stems from previous studies demonstrating that lifelong bilingualism changes WM brain structure and preserves its integrity in older adults. Our findings expand on this notion by suggesting that similar changes can be observed when learning a foreign language occurs later in life. Consistent with this, our whole-brain DTI analysis revealed improvements in axial diffusivity (AD) following four months of foreign-language training (**Fig. 2**). Positive changes in DTI parameters indicate enhanced integrity of axonal fibers. This finding aligns with a previous study by Anderson et al. (2018), which compared monolinguals and bilinguals and found greater AD in the left superior longitudinal fasciculus (lSLF), a white matter tract connecting Broca’s area with the temporal lobes, in bilingual individuals. Moreover, Luk et al. (2011) and Pliatsikas et al. (2015) reported higher fractional anisotropy (FA) in older bilinguals compared to monolinguals, and this has also been observed in younger adults. Although we did not observe an increase in FA in our study, this is not surprising given the short duration of foreign language exposure compared to lifelong bilingualism in previous investigations.

Furthermore, we observed a negative correlation between radial diffusivity (RD) and FA, suggesting that RD likely contributed to the lack of FA changes observed in our study. It is important to note that the four-month duration of our language course may not have been sufficient to induce significant alterations in FA. Nevertheless, our results provide evidence of structural changes in WM integrity following foreign language learning, supporting the notion that language learning in later life can promote neuroplasticity-related changes.

The present study is not without limitations, and it is important to acknowledge them. One noteworthy limitation is that our cognitive and structural analysis focused solely on the intervention group due to technical constraints. Sharing MRI slots with other research groups necessitated a shortened acquisition protocol. However, our comparative analysis within the intervention group before and after the intervention remains valid. The longitudinal design of the study allowed us to examine brain-behavior correlations from the beginning to the end of the language course, revealing consistent associations between Stroop task performance and changes in axial diffusivity (AD). Furthermore, our previous work demonstrated increased functional connectivity in the intervention group but not in the control group (Bubbico et al., 2019).

Another limitation is inherent to the design of non-pharmacological studies. The conventional approaches used to minimize biased observations, such as single or double-blind methodologies, cannot be easily applied in these settings. To address this, future studies could explore the dose-response effect of the training by considering factors such as the number of lessons, hours per week, duration of the course, and quantitative measures of learning abilities. Additionally, our study had a relatively small sample size, mainly due to technical and logistical challenges in organizing and managing courses for a larger audience. This limitation may impact the overall quality of the teaching experience and compliance (Campbell, 2007; Hutchins et al., 2015). Future studies examining the effects of foreign language learning in larger cohorts should also consider potential confounding factors, including the Hawthorne effect, sex-related differences, concurrent medication use, and additional comorbidity factors (McCambridge et al., 2014a, 2014b).

Furthermore, the study focused on a specific language (English) and beginner-level proficiency, which may limit the generalizability of the findings to other languages and proficiency levels. Future research should explore different languages and proficiency levels to determine if similar effects are observed. Moreover, it is important to note that the primary intention of this study was to facilitate language learning within a relatively short training duration, rather than the establishment of bilingual proficiency. Participants were engaged in a structured language learning program, but the study did not aim to achieve bilingualism within the limited intervention period. Nevertheless, the potential implications of these findings for future research in the broader context of language acquisition and brain plasticity remain relevant. Future studies that include participants with a well-established language background will be essential for further exploration in this field.

Despite these limitations, our findings provide valuable longitudinal data in older adults and contribute to our understanding of how foreign language learning affects neuroplasticity. They also raise intriguing questions for further investigation. For instance, the duration of cognitive and structural changes triggered by language learning and whether they can provide long-lasting neuroprotection similar to lifelong bilingualism warrant exploration. Additionally, investigating whether these effects can be observed in individuals with early signs of cognitive decline (such as subjective cognitive decline or mild cognitive impairment) who still exhibit levels of neuroplasticity would be valuable. A precision medicine approach could also shed light on whether certain individuals are more likely to benefit from these interventions, and whether the intervention induces biological changes that can be monitored through peripheral biomarkers. Addressing these questions in large, well-controlled cohorts could provide crucial information for implementing language learning interventions in rehabilitation and occupational settings. The present findings serve as a steppingstone toward exploring these questions and expanding our knowledge in the field.

## Conclusions

In conclusion, this study provides valuable insights into the effects of a four-month foreign language learning program on cognitive and structural changes in the brains of healthy older adults. The findings demonstrate that language learning can lead to significant increases in diffusivity measures, particularly in fronto-occipital fasciculus, the superior longitudinal fasciculus, and the corpus callosum areas. These structural changes are accompanied by improvements in executive functions, as evidenced by the positive association between changes in axial diffusivity and performance on the Stroop task.

The results support the notion that short-term language learning interventions can induce neuroplastic changes in the aging brain, specifically impacting white matter integrity. This highlights the potential of foreign language learning as a cognitive intervention to promote brain health and enhance cognitive abilities in older individuals. The present study provides evidence for the potential neuroprotective effects of language learning, suggesting that it may be an effective strategy to delay or mitigate age-related cognitive decline. Healthcare providers and policymakers could therefore incorporate language learning interventions into holistic approaches for supporting healthy aging.

These findings contribute to the growing body of research on bilingualism and its cognitive benefits, shedding light on the early stages of foreign-language acquisition in older adults. Further studies with larger sample sizes and longitudinal designs are warranted to better understand the duration and persistence of these cognitive and structural changes induced by language learning.

In summary, this study underscores the importance of foreign language learning as a potential tool for cognitive enhancement and neuroprotection in aging populations. It offers promising implications for the development of targeted interventions aimed at promoting healthy brain aging and improving cognitive functioning in older adults and support the importance of incorporating foreign language learning programs into educational curricula for older adults. Governments and educational institutions may consider implementing language learning initiatives tailored to the aging population to harness the cognitive benefits described in the study. Encouraging language learning in older adults can also foster cultural awareness and integration. Learning a new language can facilitate communication and understanding between different cultural groups, promoting social cohesion and inclusivity.

## Future works

Future studies should persist in investigating the enduring impact of language learning on brain structure and cognitive function in older adults. This research can be expanded in several directions to provide a more comprehensive understanding of the underlying mechanisms and potential benefits of language learning interventions. Firstly, larger sample sizes and control groups should be included to strengthen the statistical power and validity of the findings. Comparisons between different language learning programs or proficiency levels will help identify optimal training strategies for maximizing cognitive benefits and structural changes. Additionally, longitudinal studies are needed to examine the durability of the observed effects. Tracking participants over an extended period will provide insights into the persistence of structural changes and cognitive improvements beyond the initial language learning period. Furthermore, investigations into the relationship between language learning and specific cognitive domains, such as different kind of memory abilities, or attention, can provide a more nuanced understanding of the cognitive benefits conferred by foreign language acquisition. Integrating more advanced neuroimaging techniques beyond diffusion tensor imaging (DTI), such as functional magnetic resonance imaging (fMRI), can provide a more comprehensive evaluation of the neural alterations linked to foreign language acquisition. Examining functional connectivity patterns and brain network dynamics in a larger sample, may reveal additional insights into the mechanisms underlying the observed structural changes. Furthermore, taking into account individual differences, including the subject’s age at the time of acquisition, language proficiency, and genetic factors, can aid in identifying the factors that contribute to variations in cognitive and structural responses to language learning. In conclusion, future research should aim to build upon the present study’s findings by investigating the long-term effects of language learning, exploring specific cognitive domains, incorporating advanced neuroimaging techniques, and considering individual differences. These efforts will contribute to a more comprehensive understanding of the potential benefits of foreign language learning interventions and their implications for promoting cognitive health in older adults.

## Ethics statement

This study was conducted in accordance with the principles outlined in the Declaration of Helsinki. Prior to participation, all subjects provided written informed consent. The research protocol received approval from the Ethics Committee of G. d’Annunzio University of Chieti-Pescara, Italy, ensuring adherence to ethical standards and protection of participants’ rights.

## Credit author statement

The idea for the study was conceived by GB. GB and MGP designed the research protocol. GB, AG, and JGG contributed to the initial drafting of the manuscript. GB, CC, MB, and MF provided supervision throughout the experiments, data exportation, and manuscript review. RN and PC conducted the TBSS analysis. JGG, RN, ASC, and GB performed the statistical analysis. MGP and AF provided research supervision and reviewed the manuscript for intellectual content. RN and ASC reviewed the manuscript for methodological rigor regarding the DTI procedures. AT conducted neuroradiological examinations. All authors have reviewed and approved the final version of the manuscript.

## Data Availability

All data produced in the present study are available upon reasonable request to the authors.

## Acknowledgement

We express our gratitude to the dedicated MRI technicians, D. Calvo Garcia and D. Petrucci, for their invaluable support during subject acquisition. We extend our sincere appreciation to all study participants for their time and commitment. We would like to thank Prof. N. Ciacio and Prof. F. D’Ettorre for their contributions to conducting language lessons at the C. d’Ascanio College of Montesilvano. We are also grateful to Mr. G. Tini and the Mazzaferro University of Third Age of Montesilvano for their assistance in subject recruitment and organization of the language lessons. We extend our sincere gratitude to Stefano Delli Pizzi for his invaluable support and guidance in overseeing the analysis of white matter tracts.

